# Easing COVID-19 lockdown measures while protecting the older restricts the deaths to the level of the full lockdown

**DOI:** 10.1101/2020.05.08.20095380

**Authors:** A.S. Fokas, J. Cuevas-Maraver, P.G. Kevrekidis

## Abstract

Guided by a rigorous mathematical result, we have earlier introduced a numerical algorithm, which using as input the cumulative number of deaths caused by COVID-19, can estimate the effect of easing of the lockdown conditions. Applying this algorithm to data from Greece, we extend it to the case of two subpopulations, namely, those consisting of individuals below and above 40 years of age. After supplementing the Greek data for deaths with the data for the number of individuals reported to be infected by SARS-CoV-2, we estimated the effect on deaths and infections in the case that the easing of the lockdown measures is different for these two subpopulations. We found that if the lockdown measures are partially eased only for the young subpopulation, then the effect on deaths and infections is small. However, if the easing is substantial for the older population, this effect may be catastrophic.

## Introduction

SARS-CoV-2, the causative virus of the current pandemic, has 75-80% identical viral genome sequence with the coronaviruses (MERS)-CoV and SARS-CoV^1^. Fortunately, it has lower mortality rates than these two viruses which caused outbreaks in 2012 and 2002^2^. However, SARS-CoV-2 is highly contagious. Perhaps the increased transmissivity of SARS-CoV-2 is due to its dual receptor attachment in the host cells: it has been shown that the attachment of SARS-CoV-2 to the surface of respiratory cells is mediated by certain viral proteins which bind not only to the angiotensin converting enzyme-2 (ACE-2) receptor^3^, but also bind to sialic acid containing glycoproteins and gangliosides that reside on cell surfaces^4^ (this is to be contrasted with SARS-CoV that binds only to ACE-2 receptors^3–5^). As a result of this high transmissivity, as well as of the delay of the international community to take appropriate preventive measures, SARS-CoV-2 has caused a pandemic, which represents the most serious global public health threat since the devastating 1918 H1N1 influenza pandemic that killed approximately 50 million people (in proportion to today’s population, this would correspond to about 200 million people).

In order to combat this threat, several countries used justifiably draconian measures which culminated in a complete lockdown. As a result of these measures, the curves depicting the total persons infected by SARS-CoV-2, in China, South Korea, many European countries, USA, and several other countries, passed by April 2020 the inflection point, namely the point where the rate of infected individuals reached its maximum. For the above countries, the curve depicting the number of deaths as a function of time also past the inflection point. Remarkably accurate predictions for the time that a plateau would be attained (defined as the time when the rate of deaths is 5% of the maximum rate) as well as the corresponding number of total deaths at that time, were presented, e.g., in^6^. Similar results regarding reported infections were presented in^7^). In addition to these works, a great variety of models have appeared in the literature stemming from deterministic, as well as probabilistic variations of the broad SIR class of models. These models aim to capture the epidemics of different countries^8–11^, or states as well as provinces^12–17^. These papers are only indicative examples of an ever-expanding volume of literature of well over 5000 papers within the first few months of the pandemic alone, in arXiv, medRxiv and bioRxiv.

Following the expected decline in several European countries of the ‘first wave’ of infections, the lockdown measures began to be lifted in May. The importance of easing the lockdown measures differently, depending on the age of individuals, is argued in^18^. This was considered vital, not only for economical but also for health considerations. Indeed, the psychological impact on the population at large of the imposed restrictions was substantial^19^. Furthermore, such features are expected to worsen, especially due to the effect of the post-traumatic disorder.

Naturally, the best possible scenario of minimizing the possible impact of lifting the lockdown measures and to prevent new waves would be the availability of a safe and effective vaccine. However, it appears that this will not be broadly available still for some time. The second optimal solution would be the discovery of effective pharmacological interventions. In this direction unprecedented efforts are underway, it is noted that the Federal Drug Administration of USA has granted a conditional approval to the anti-viral medication Remdesivir^20^. Also, dexamethasone reduced mortality among hospitalized patients who received supplemental oxygen and the benefit was greatest among patient who underwent mechanical ventilation^21^. Furthermore, including the testing of old or new medications as well as the employment of specific monoclonal antibodies as bamlanivimab (LY-CoV555)^22^, whose conditional approval has also been granted by the FDA. For example, there are ongoing clinical trials testing the synthetic protein tocilizumab that binds interleukin-6 (often used in rheumatoid arthritis), as well as the infusion of COVID-19 convalescent plasma^23^. Unfortunately, the combination of the anti-viral medications lopinavir and ritonavir that are effective against the human immunodeficiency virus has not shown to be beneficial^24^; similarly, the combination of the anti-malarial medication hydroxychloroquine and the antibiotic azithromycin, is not only ineffective but can be harmful^25,26^.

Taking into consideration that a decision was made to ease the lockdown measures without the benefit of a proven pharmacological cover against the multitude of possible ways that SARS-CoV-2 can attack the human body^27^, the following natural questions arise: is it possible to predict the effect on the number of infected and the number of deaths following the easing of the lockdown measures? If the answer to this question is positive, is it possible to use this insight for designing an optimal exit strategy? The main goal of this paper is to address these questions for the COVID-19 pandemic in Greece.

The issue of easing the lockdowns is one that has widely concerned both politicians and the scientific community; see, e.g.,^28^ for an example analysis on Italy performed by the widely recognized group at Imperial College. On the one hand, there have been calls for such easing accompanied by increased testing, broader communication and other similar measures^29^. Other studies^30^ have focused their efforts on the examination of effects to the global supply-chains and have recommended a cautious approach as more beneficial to the global economy. Additional work has considered this ending of lockdown as an optimization process considering “on-off” approaches partially or fully within the population^31^. Identifying a gap in the literature regarding the examination of different age groups and strategies associated with them, our interest herein is to explore different scenarios at the end of the pandemic’s first wave towards easing the lockdown measures. Leveraging a suitable variation of the well-known compartmental models in epidemiology^32,33^ properly adapted to the special features of SARS-CoV-2 (such as, e.g., the role of asymptomatic virus carriers), we identify optimal model parameters for both single- and two-age populations. Subsequently, we utilize this knowledge to test different scenarios of lockdown release, and unveil conditions that may be tolerable (such as the partial release of the young population through a modest increase in contacts) vs. ones that are catastrophic (such as the release of the full population).

## Results

### Overview

In earlier work^34^, epidemiological models are broadly divided into two large categories, called *forecasting* and *mechanistic*. The former models fit a specific curve to the data and then attempt to predict the dynamics of the quantity under consideration. The most well known mechanistic models are the SIR-type models. As noted by Holmadahl and Buckee^34^, the mechanistic models involve substantially more complicated mathematical machinary than the forecasting models, but they have the advantage that they can make predictions even when the relevant circumstances change. In our case, since our goal is to make predictions after the situation changes due to the lifting of the lockdown measures, we need to consider a mechanistic model. However, it is widely known that the main limitation of mechanistic models is the difficulty of determining the parameters specifying such models. In this direction, a methodological advance was presented by the authors^35^, filling an important gap in the relevant literature: it was shown in this work that from the knowledge of the most reliable data of the epidemic in a given country, namely the cumulative number of deaths, it is possible to determine suitable combinations of the constant parameters (of the original model) which specify the differential equation characterizing the death dynamics. Furthermore, a robust numerical algorithm was presented for obtaining these parameters. One of these constants, denoted by *c*, is particularly important for the analysis of the effect of easing the lockdown conditions, because it is proportional to the number of contacts between asymptomatic individuals that are infected by SARS-CoV-2 and susceptible ones. More concretely, as the equations presented below will indicate, this coefficient is measured in units of inverse population (where the population represents the number of individuals to which we assign no units) times inverse days and reflects not only the number of contacts, but also the probability of infection given a contact which is proportional to the viral load (i.e., the viral concentration in the respiratory-tract fluid) of expelled respiratory droplets^36^. Easing the lockdown will lead to an increase of the value of this constant. Thus, in order to quantify this effect we assumed that the post-lockdown situation could be described by the same model but with *c* multiplied by an integer number *ζ*, such as *ζ* = 2, or 3, etc. Notice that assuming a fixed viral load emission (i.e., no face mask or similar protective measures), this would be tantamount to doubling or tripling the number of contacts per day. To put things in perspective, it is relevant to mention here that in the relevant literature a ballpark estimate for daily contacts of an individual is about 13.4^37^.

We first applied the above algorithm to the case of the COVID-19 epidemic in Greece. However, the novelty and the main interest of the present work consists of the extension and application of the above methodology to two subpopulations. This situation is significantly more complicated than that of^2^ and is described by 12 ODEs involving 18 parameters (details are discussed in the Methods section). Using this extended formulation, we analysed the effect of easing the lockdown measures under two distinct possible scenarios: in the first, we examined what would happen if the interactions between older persons, namely persons above 40 years of age, as well as between older and younger persons, namely those below 40, continue to be dictated by the same restrictions as those of the lockdown period. However, we assumed that the interaction among the young was progressively more free. In the second case, we analysed the effect of easing the lockdown measures in the entire population without distinguishing the older from the young. In principle, the effect on deaths in the above two scenarios could be analyzed by the extension of the rigorous results of^35^. However, due to the sparsity of the deaths data (especially for the younger population), this approach is practically not possible at present. Thus, we supplemented the data for deaths for the two subpopulations with data for the cumulative numbers of reported infected.

Using four sets of data, namely the number of deaths and the number of reported infected for the older and the younger population we found that the above two alternatives would result in very different outcomes: in the first case, the total number of deaths of the two sub-populations and the number of total infections would be relatively small. In the second case, these numbers would be prohibitively high. Specifically, in the case of Greece, if the lockdown was to be continued indefinitely, our analysis suggests that the total numbers of deaths and infections would finally be around 165 and 2550, respectively. These numbers would remain essentially the same even if the lockdown measures for the interaction between the young people were eased substantially, provided that the interactions of older-older and older-young would remain the same as during the lockdown period. For example, even if the parameter measuring the effect of the lockdown restrictions on the young-young interactions were increased fourfold, the number of deaths and infections would be (according to the model extrapolation) 184 and 3585, respectively. On the other hand, even if the parameters characterizing all three interactions were increased only threefold, the relevant numbers would be 48144 and 1283462. It is clear that the latter numbers are prohibitive, suggesting that a generic release of the lockdown may be catastrophic.

In our view, the explanations provided in the Methods Section for the assumptions of our model, which show that these assumptions are typical in the standard epidemiological models, substantiate the qualitative conclusions (and notes of caution) regarding the impact of the above two different types of exit policies. This may provide a sense of how a partial restoration of regular life activities can be achieved without catastrophic consequences, while the race for pharmacological or vaccine-based interventions that will lead to an end of the current pandemic is still ongoing. Importantly, we also offer some caveats emphasizing the qualitative nature of our conclusions and possible factors that may substantially affect the actual outcome of the lifting of lockdown measures.

### Model Setup: Single Population vs. Two Age Groups

We divide the population in two subpopulations, the young (y) and the older (o). In order to explain the basic assumptions of our model we first consider a single population, and then discuss the needed modifications in our case which involves two subpopulations. Let *E*(*t*) denote the exposed (but not infectious) population. An individual in this population, after a median 4-day period (required for incubation — see e.g.^38^) will either become sick or will be asymptomatic; an interval of 3-10 days captures 98% of the cases. The sick (infected) and asymptomatic populations will be denoted, respectively, by *I*(*t*) and *A*(*t*). The rate at which an exposed person becomes asymptomatic is denoted by *a*; this means that each day *aE*(*t*) persons leave the exposed population and enter the asymptomatic population. Similarly, each day *sE*(*t*) leave the exposed population and enter the sick population. These processes, as well as the subsequent movements are depicted in the flowchart of Fig. 1.

**Figure 1.**
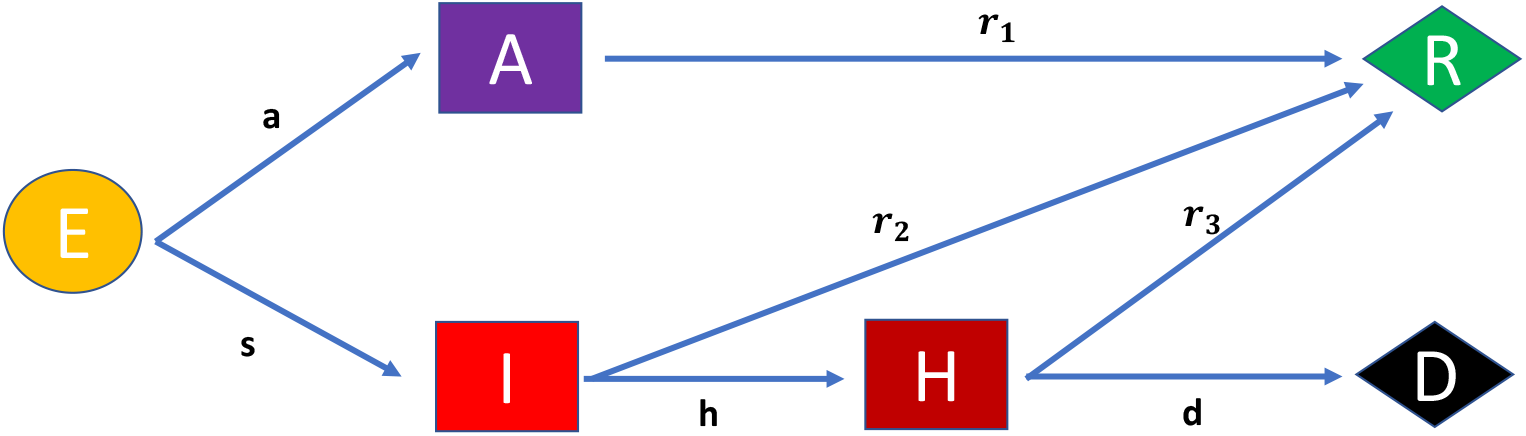
Flowchart of the populations considered in the model and the rates of transformation between them. The corresponding dynamical equations are Eqs. (1)–(6).

The asymptomatic individuals recover with a rate *r*_1_, i.e., each day *r*_1_*A*(*t*) leave the asymptomatic population and enter the recovered population, which is denoted by *R*(*t*). The sick individuals either recover with a rate *r*_2_ or they become hospitalized, *H*(*t*), with a rate *h*. In turn, the hospitalized patients also have two possible destinations; either they recover with a rate *r*_3_, or they become deceased, *D*(*t*), with a rate *d*.

It is straightforward to write the above statements in the language of mathematics; this gives rise to the equations (1)–(5) below:

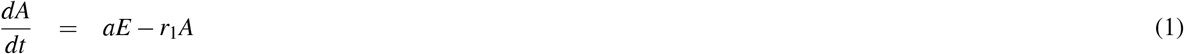

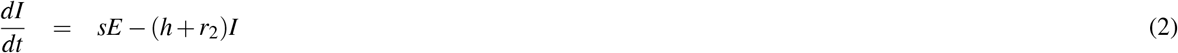

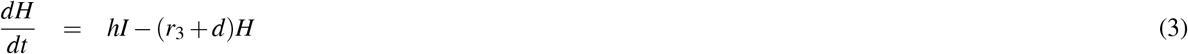

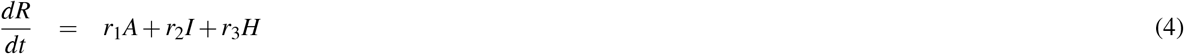

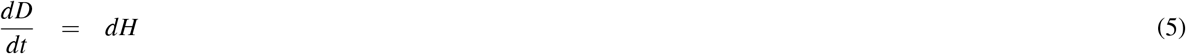

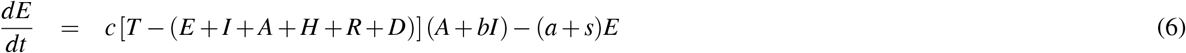

It is noted that our model is inspired by various expanded versions of the classic SIR model adapted to the particularities of COVID-19 (such as the key role of the asymptomatically infected). It is, in particular, inspired by, yet not identical with that of^14^. In order to complete the system of equations (1)–(6), it is necessary to describe the mechanism via which a person can become infected. For this purpose we adopt the standard assumptions made in the typical epidemiological models, such as the SIR (susceptible, infected, recovered) model: let *T* denote the total population and let *c* characterize the number of contacts per day made by an individual with the capacity to infect (*c* is thought of as being normalized by *T)*. Such a person belongs to *I, A* or *H*. However, for simplicity we assume that the hospitalized population *cannot* infect; this assumption is based on two considerations: first, the strict protective measures taken at the hospital, and second, the fact that hospitalized patients are infectious only for part of their stay in the hospital. The latter fact is a consequence of the relevant time scales of virus shedding in comparison to the time to hospitalization and the duration of hospital stay. The asymptomatic individuals are (more) free to interact with others, whereas the (self-isolating) sick persons are not. Thus, we use *c* to characterize the contacts of the asymptomatic persons and *b* to indicate the different infectiousness (due to reduced contacts/self-isolation) of the sick in comparison to the asymptomatic individuals.

The number of people available to be infected (i.e., the susceptible population) is *T −* (*E* + *I* + *A* + *H* + *R* + *D*). Indeed, the susceptible individuals consist of the total population minus all the individuals that are going or have gone through the course of some phase of infection, namely they either bear the infection at present (*E* + *A* + *I* + *H*) or have died from COVID-19 (*D*) or are assumed to have developed immunity to COVID-19 due to recovery (*R*). Hence, if we call the total initial individuals *T*, this susceptible population is given by the expression written earlier. The rate by which each day individuals enter *E* is given by the product of the above expression with *c*(*A* + *bI*). At the same time, as discussed earlier, every day (*a* + *s*)*E* persons leave the exposed population. It is relevant to note here that within this simpler model, it is possible to calculate the basic reproduction number *R*_0_, which is a quantity of substantial value in epidemiological studies^32,33^. In this model, this can be found to be^33^:

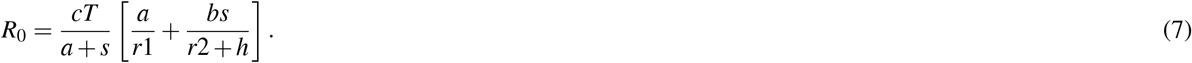

This will be useful below for the purposes of finding the change in *c* (under lockdown) needed in order for transmission to cross the threshold of *R*_0_ = 1 and thus to lead to growth of the epidemic. In the particular case of the data shown in Table 1, *R*_0_ = 0.4084, in accordance with the lockdown situation associated with a controlled epidemic.

**Table 1.**
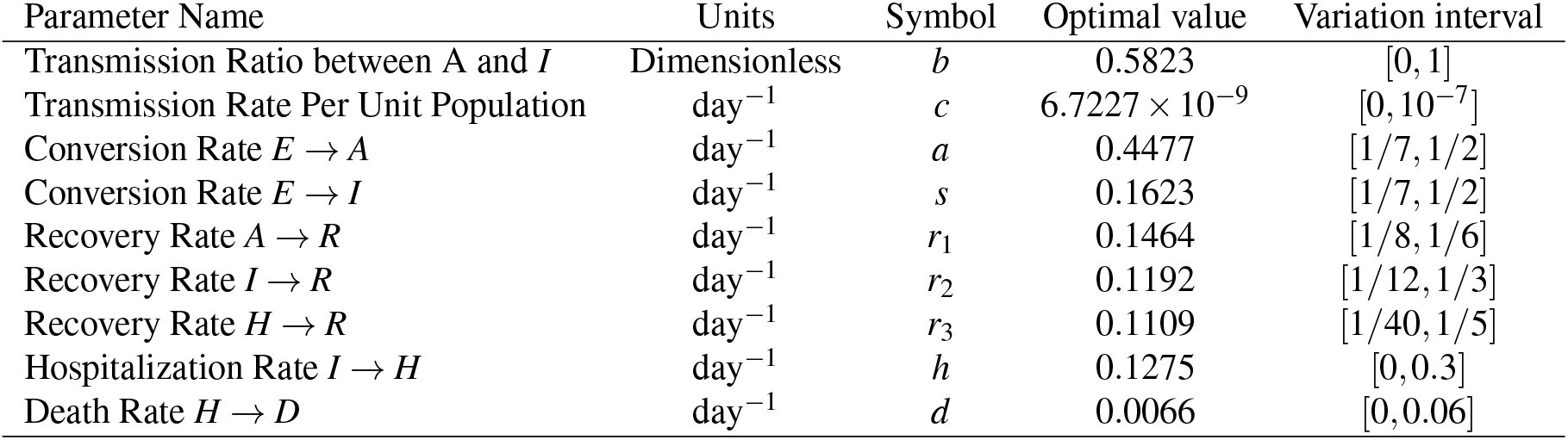
Optimized Model Parameters for the single population model, and the variation interval of each parameter within the optimization process (for further details, see Methods Section).

| Parameter Name | Units | Symbol | Optimal value | Variation interval |
| --- | --- | --- | --- | --- |
| Transmission Ratio between A and I | Dimensionless | $b$ | 0.5823 | [0, 1] |
| Transmission Rate Per Unit Population | $\text{day}^{-1}$ | $c$ | $6.7227 \times 10^{-9}$ | $[0, 10^{-7}]$ |
| Conversion Rate $E \rightarrow A$ | $\text{day}^{-1}$ | $a$ | 0.4477 | $[1/7, 1/2]$ |
| Conversion Rate $E \rightarrow I$ | $\text{day}^{-1}$ | $s$ | 0.1623 | $[1/7, 1/2]$ |
| Recovery Rate $A \rightarrow R$ | $\text{day}^{-1}$ | $r_1$ | 0.1464 | $[1/8, 1/6]$ |
| Recovery Rate $I \rightarrow R$ | $\text{day}^{-1}$ | $r_2$ | 0.1192 | $[1/12, 1/3]$ |
| Recovery Rate $H \rightarrow R$ | $\text{day}^{-1}$ | $r_3$ | 0.1109 | $[1/40, 1/5]$ |
| Hospitalization Rate $I \rightarrow H$ | $\text{day}^{-1}$ | $h$ | 0.1275 | [0, 0.3] |
| Death Rate $H \rightarrow D$ | $\text{day}^{-1}$ | $d$ | 0.0066 | $[0, 0.06]$ |

It is straightforward to modify the above model so that it can describe the dynamics of the older and younger subpopulations. Each subpopulation satisfies the same set of equations as those described above, except for the last equation which is modified as follows: the people available to be infected in each subpopulation are described by the expression given above where *T, E, I, A, H, R, D* have the superscripts ^*o*^ or ^*y*^, denoting older and young, respectively; *A* + *bI* is replaced in both cases by *A*^*o*^ + *A*^*y*^ + *b*(*I*^*o*^ + *I*^*y*^) where for simplicity we have assumed that the infectiousness of the older and the young is the same. We have already considered the implications of the generalisation of the above model by allowing different parameters to describe the interaction of the older and young populations; this will be discussed in the Methods Section. In what follows, we will discuss the results of this simpler “isotropic” interaction model.

### Quantitative Model Findings

The parameters of the model are given in the flowchart of Fig. 1. Naturally, for the two-age model considered below, there is one set of such parameters associated with the younger population and one associated with the older one. The optimization routine used for the identification of these parameters is explained in detail in the Methods Section. The parameters resulting from this optimization for the single population model are shown in Table 1, whereas for each of the two populations are given in Table 2. Clearly, many of these parameters are larger for the older population in comparison to the young, leading to a larger number of both infections and deaths in the older than in the young population.

**Table 2.** Optimized (Isotropic) Model Parameters for the young and older populations, and the variation interval of each parameter within the optimization process (for further details, see Methods Section).

| Parameter Name | Units | Symbol | Optimal value | Variation interval |
| --- | --- | --- | --- | --- |
| Transmission Ratio between A and I | Dimensionless | $b$ | 0.2936 | [0, 1] |
| Transmission Rate Per Unit Population | $\text{day}^{-1}$ | $c$ | $7.4941 \times 10^{-9}$ | $[0, 10^{-7}]$ |
| Conversion Rate $E \rightarrow A$ (young population) | $\text{day}^{-1}$ | $a^y$ | 0.2715 | $[1/7, 1/2]$ |
| Conversion Rate $E \rightarrow I$ (young population) | $\text{day}^{-1}$ | $s^y$ | 0.0751 | $[1/50, 1/6]$ |
| Conversion Rate $E \rightarrow A$ (older population) | $\text{day}^{-1}$ | $a^o$ | 0.2437 | $[1/35, 1/2]$ |
| Conversion Rate $E \rightarrow I$ (older population) | $\text{day}^{-1}$ | $s^o$ | 0.0846 | $[1/10, 1/3]$ |
| Recovery Rate $A \rightarrow R$ (young population) | $\text{day}^{-1}$ | $r_1^y$ | 0.1360 | $[1/8, 1/6]$ |
| Recovery Rate $I \rightarrow R$ (young population) | $\text{day}^{-1}$ | $r_2^y$ | 0.1526 | $[1/12, 1/4]$ |
| Recovery Rate $H \rightarrow R$ (young population) | $\text{day}^{-1}$ | $r_3^y$ | 0.1094 | $[1/20, 1/5]$ |
| Recovery Rate $A \rightarrow R$ (older population) | $\text{day}^{-1}$ | $r_1^o$ | 0.1343 | $[1/8, 1/6]$ |
| Recovery Rate $I \rightarrow R$ (older population) | $\text{day}^{-1}$ | $r_2^o$ | 0.2668 | $[1/20, 1/3]$ |
| Recovery Rate $H \rightarrow R$ (older population) | $\text{day}^{-1}$ | $r_3^o$ | 0.0801 | $[1/30, 1/5]$ |
| Hospitalization Rate $I \rightarrow H$ (young population) | $\text{day}^{-1}$ | $h^y$ | 0.0173 | [0, 0.1] |
| Death Rate $H \rightarrow D$ (young population) | $\text{day}^{-1}$ | $d^y$ | 0.0017 | [0, 0.05] |
| Hospitalization Rate $I \rightarrow H$ (older population) | $\text{day}^{-1}$ | $h^o$ | 0.3120 | [0.1, 0.5] |
| Death Rate $H \rightarrow D$ (older population) | $\text{day}^{-1}$ | $d^o$ | 0.0103 | [0, 0.2] |

Support for the validity of our model is presented in Fig. 2, which depicts its comparison (using the above optimized parameters) with the available data. The situation corresponding to keeping the lockdown conditions indefinitely, is the one illustrated in Fig. 2. In this case, the number of deaths and cumulative infections rapidly reaches a plateau, indicating the elimination of the infection. Here, we have optimized the model on the basis of data used from Greece^39^ between April 3rd and May 4th. It is noted that daily updates occur at 3pm for the country of Greece, hence it is not clear up to what time the data are collected that are included in the daily report. We have assumed that the data reflect the infections and deaths present on that particular day. This possibly shifts the starting point of our count by a few hours, but should not change the overall result trends.

**Figure 2.**
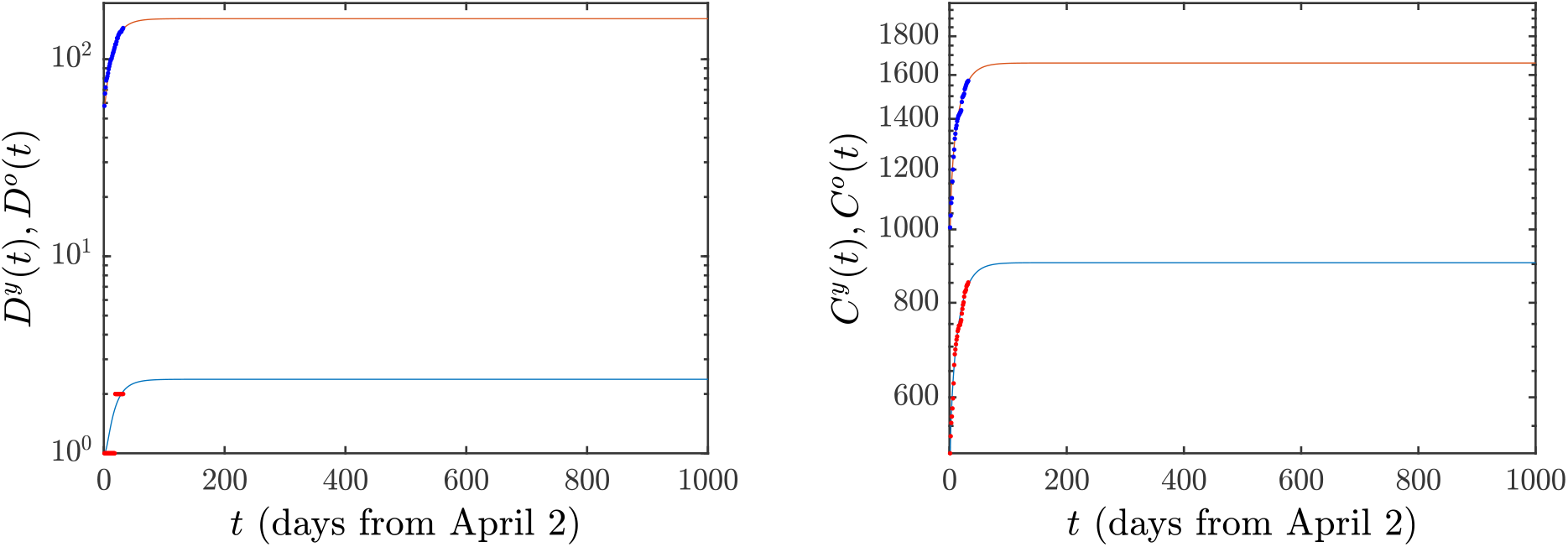
Evolution of the current situation of deaths *D*(*t*) (left) and cumulative infections *C*(*t*) (right) in Greece, under the case of an indefinite continuation of the lockdown conditions. In this and all the figures that follow, the blue curve corresponds to the young population, while the red curve to the older population. The data for Greece from the 3rd of April to the 4th of May 2020 are depicted by dots. For the latter, alternate colors have been used (i.e., blue dots for the older population and red for the younger for clearer visualization).

We next explain the implications of the model when different scenarios of ‘exit’ from the lockdown state are implemented. The relevant results are illustrated in Figs. 3-4 and the essential conclusions are summarized in Table 3 for the numbers of deaths and cumulative infections, respectively. First, we need to explain the meaning of the parameter *ζ* appearing in the above tables: this parameter reflects the magnitude of the easing of the lockdown restrictions. Indeed, since the main effect of the lessening of these restrictions is that the number of contacts increases, we model the effect of easing the lockdown restrictions by multiplying the parameter *c* with a factor that we refer to as *ζ*. The complete lockdown situation corresponds to *ζ* =1; the larger the value of *ζ*, the lesser the restrictions imposed on the population. By employing the above quantitative measure of easing the lockdown restrictions, we consider in detail two distinct scenarios. In the first, which corresponds to the top rows of the Figures 3 and 4, we only allow the number of contacts of “young individuals with young individuals” (corresponding to the parameter *c*^*yy*^ mentioned in the Methods Section) to be multiplied by the factor *ζ*. This means that the lockdown measures are eased only with respect to the interaction of young individuals with other young individuals, while the interactions of the young individuals with the older ones, as well as the interactions among older individuals remain in the lockdown state. In the second scenario, corresponding to the bottom rows of the Figures 3 and 4, the restrictions of the lockdown are simultaneously eased in both the young and the older population; in this case all contacts are increased by the factor *ζ*. It is noted that while we change *c* by this factor, we maintain the product *cb* at its previous value (i.e., we concurrently transform *c → ζc* and *b→ b/ζ)* considering that the sick still operate under self-isolation conditions and thus do not accordingly increase their number of contacts.

**Table 3.** Deaths *D*(*t*) and Cumulative infections *C*(*t*) in the case of increasing of the number of contacts by *ζ*. The second and fourth columns refer to the case for which the lockdown measures are eased for the young population, whereas the third and fifth column refer to the one where this occurs for both the young and older populations.

| $\zeta$ | Deaths | | Cumulative infections | |
| --- | --- | --- | --- | --- |
|  | Y Only Release | Y+O Release | Y Only Release | Y+O Release |
| 1 | 164 | 164 | 2564 | 2564 |
| 2 | 165 | 222 | 2622 | 4114 |
| 3 | 167 | 48144 | 2762 | 1283462 |
| 4 | 184 | 72180 | 3585 | 1925122 |
| 5 | 6044 | 83274 | 306219 | 2221296 |

**Figure 3.**
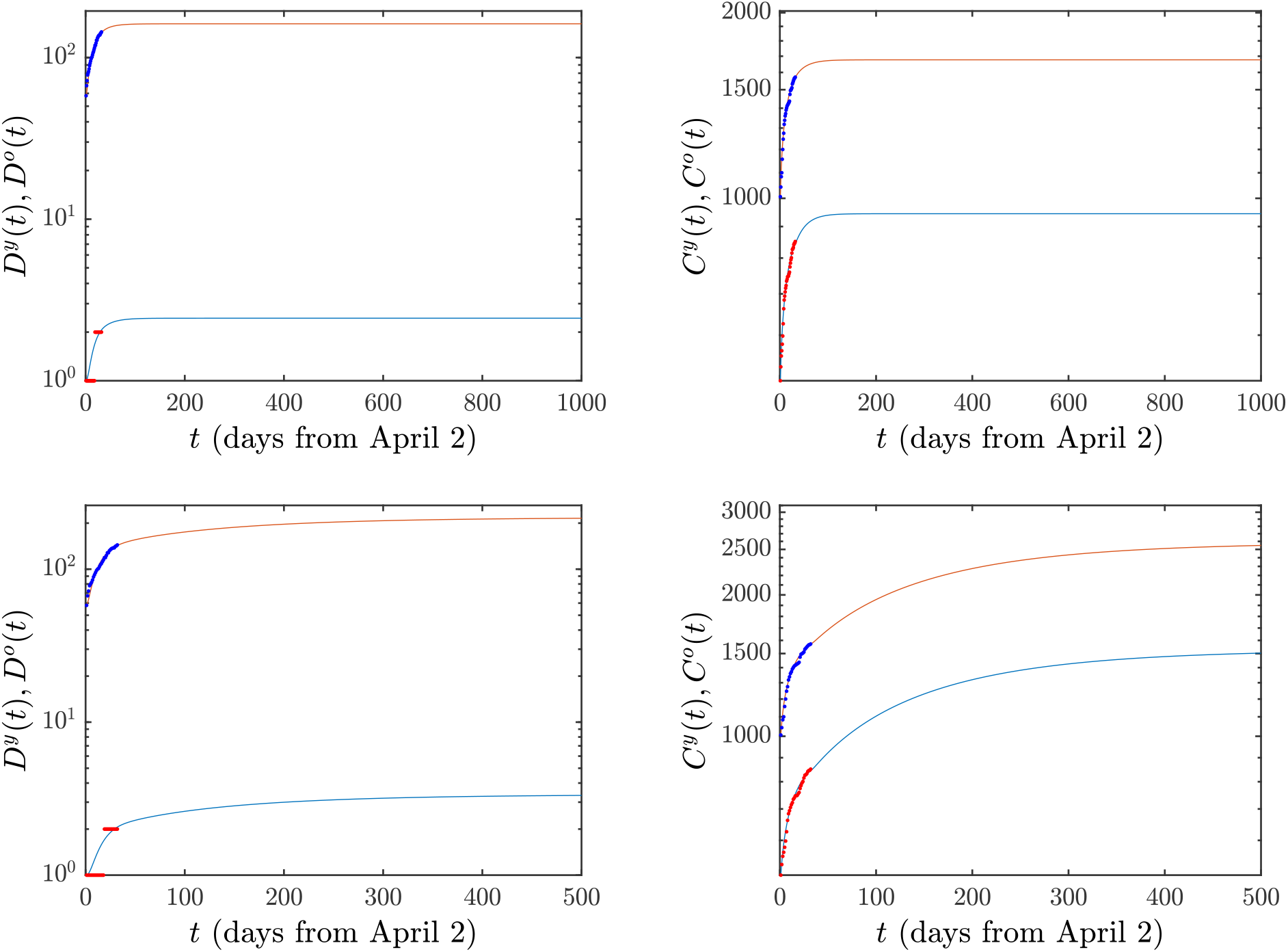
Again the Deaths *D*(*t*) and the cumulative infections *C*(*t*) are given for the case where the *c* factor (characterizing the number of contacts) amongst young individuals is doubled, but those of the older individuals (and of the young-older interaction) are kept fixed. This is shown in the top panels. In the bottom panels, the *c*’s of both young and old individuals are doubled.

**Figure 4.**
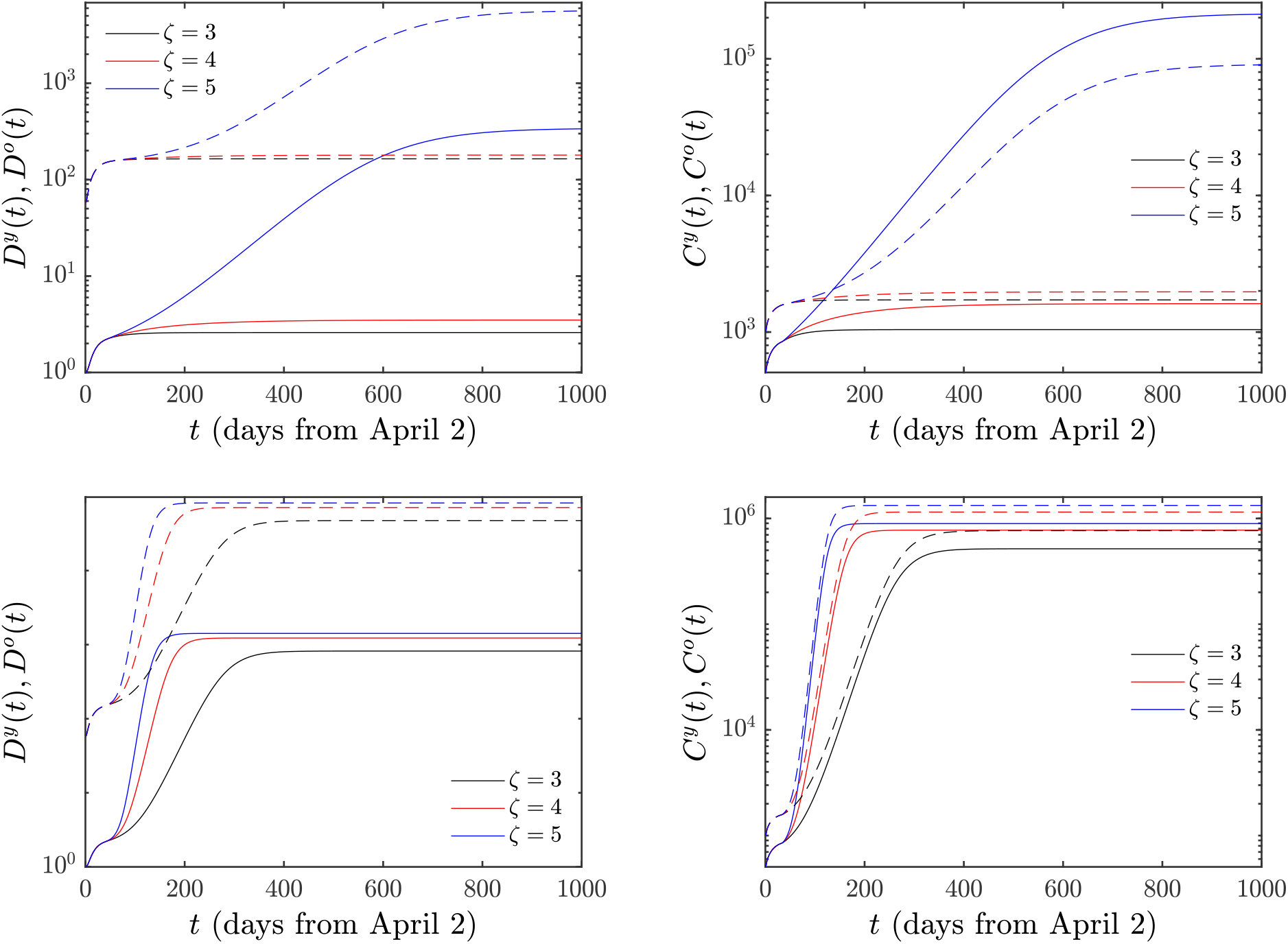
Same as reported in Fig. 3 but now where the contacts are multiplied by factors 3, 4 and 5. Full (dashed) lines hold for the young (older) population.

Fig. 3 corresponds to the case where the parameter *ζ* associated with the number of contacts between susceptible and asymptomatic individuals *doubles*. In this case, as also shown in Table 3, the situation does not worsen in a dramatic way. In particular, the number of deaths increases by 1, whereas the cumulative infections only increase by the small number of 58. In the second scenario where the number of contacts is doubled for both the young and the older populations, we find slightly larger (but not totally catastrophic) effects: the number of deceased individuals increases by 58 and the total number of infections grows by 1550.

The situation becomes far more dire when the number of contacts is multiplied by a factor of 3 for both the young and older populations, meaning that the lockdown restrictions are eased significantly for the entire population. As shown in Table 3 and in Fig. 4, if the *c*’s of the young population only are multiplied by a factor of 3, then the deaths are increased by 3 and the infections by 198 (black line in the Figure and 3rd row of the Tables). This pales by comparison to the dramatic scenario when the *c*’s associated with both the young and older sub-populations are multiplied by 3; in this case, the number of deaths jumps dramatically to 48144, while the number of infections is a staggering 1283462, growing by about 500 times.

An example corroborating the above qualitative trend can also be found in Fig. 4 and in the 4th and 5th rows of Table 3. Here, for e.g. *ζ* = 5, even the effect of releasing solely the young population leads to very substantial increases, namely to 6044 deaths and 306219 infections although of course it is nowhere near the scenarios of releasing both young and older populations. In the second scenario, the numbers are absolutely daunting: using the parameters of Table 2 we find that the number of deaths jumps to 83274 and the number of cumulative infections to 2221296.

Finally, we show the prediction of the easing measures in the hospitalizations (i.e. daily occupied beds in hospitals). This is a crucial point to assess in order that the health system does not collapse because of COVID-19 patients. Figure 5 shows these trends for the above mentioned values of *ζ*. In the case of releasing solely the young population (see left panel of the Figure), it is observed that the number of hospitalizations decreases monotonically except for *ζ* = 5, where the hospitalization peak is 523 for the young population and 1426 for the older one (values that are affordable by Greek health system); however, if both the young and older population are released (see right panel of the Figure), there is a monotonically decreasing behaviour only for *ζ* = 1 and 2. For higher *ζ* we observe that the height of the peak obviously increases with *ζ*, while this peak also occurs earlier when the number of contacts is increased; for instance, for *ζ* = 3, the hospitalization peak number of the young population is 3844 whereas this value is 37030 for the older one, numbers that are, unfortunately, unaffordable for the Greek health system. These figures grow even further to 16869 and 163648 if *ζ* = 5.

**Figure 5.**
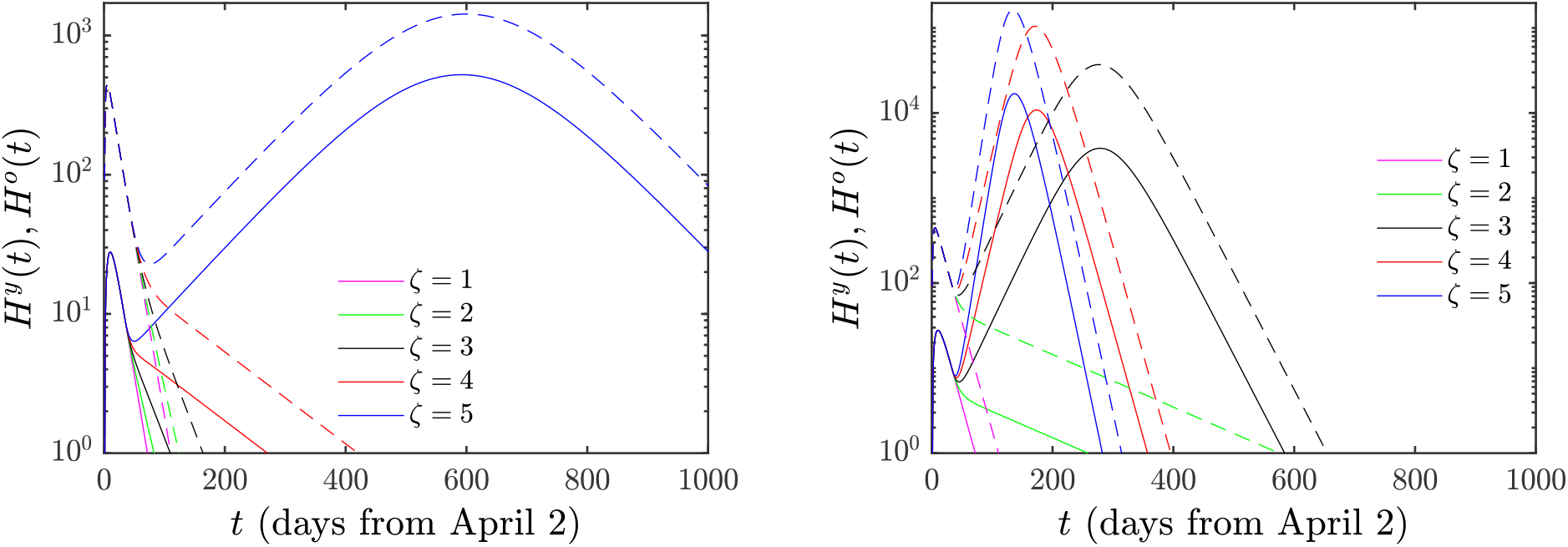
Hospitalizations when only the young population (left) or both the young and older (right) population are released. Full (dashed) lines hold for the young (older) population.

In light of the above results, the significance of preserving the lockdown restrictions of the sensitive groups of the older population is naturally emerging. It can be seen that in the case where the number of contacts is roughly doubled, the behavior of release of young or young and older individuals is not dramatic (although even in this case releasing only the young population is, of course, preferable). Nevertheless, a more substantial release of the young population is still not catastrophic. On the other hand, the higher rates of infection, hospitalization and proneness to death of senior individuals may bring about highly undesirable consequences, should both the young and older members of the population be allowed to significantly increase (by 3 times or more) their number of contacts.

## Discussion

The above results are both encouraging and at the same time raise the possibility of a potential disaster. The encouraging results are twofold: (i) If the number of contacts in the population at large is roughly doubled in comparison to the fully lockdown situation, the number of deaths and infections increases only slightly. (ii) If the older population continues to observe the conditions of the lockdown, but the young people are allowed to interact among themselves relatively freely, the number of deaths and of infected persons remains relatively small.

The first result above suggests that a mild modification of the lockdown restrictions (where the number of contacts doubles) is relatively safe, irrespectively of whether this process is applied to the young members of the population (top rows of Fig. 3) or both to the young and the older persons (bottom rows of Fig. 3). The fact that *ζ* = 2 gives rise to a stable situation is consistent with the following considerations: mathematically, the notion of stability is usually formulated in terms of the concept of the so-called “dominant eigenvalue”. For our case, by computing this eigenvalue for the *healthy state* (where no individuals are at any stage of sickness, symptomatic or asymptomatic), we find that the transition between stability and instability occurs if the dominant eigenvalue vanishes and this corresponds to a scenario where *c* equals 2.0923 times the value of *c* corresponding to the lockdown conditions. In epidemiological language, this corresponds to the scenario of *R*_0_ = 1, as the growth of the epidemic is tantamount to the largest eigenvalue of the corresponding vanishing equilibrium (healthy) state becoming larger than 0, hence reflecting its instability. Thus, the transition from stability to instability occurs between doubling and tripling of all contacts. Indeed, in this case, the analysis of the model provides a warning of a possible disaster: if the number of contacts of the older population increases threefold, either among themselves or with the young population, the increase of deaths and of the number of infected persons is dramatic. On the other hand, if only the young population is released, the critical value of the corresponding *c*^*yy*^ is 3.3848 larger than the one corresponding to lockdown conditions, so it is safe to allow the young population to even triple its contacts. Moreover, we have seen that even increasing that number of contacts by a factor of 4 does not have catastrophic consequences. More dire is the situation when the relevant factor is 5 but even then it leads to a far less significant effect than, e.g., allowing both the young and the older population to triple their contacts. A relevant, suggestive comparison to perhaps add here (yet not in any way a “definitive” one) is that in a recent, ongoing study by some of the authors in a single-age population model including both data prior to quarantine (“normal conditions”) and ones post-quarantine, the relevant ratio of *ζ* between the two was found to be between 3 and 4^40^. This suggests that in that single-age model an increase of the contacts by such a factor is proximal to a return to pre-quarantine conditions for the cases of Greece and Andalusia considered therein.

Possible weakness of our results include the following: first, there is discrepancy between the function *C*(*t*) of the total number of (truly) ‘infected’ individuals calculated in our model, and the number of ‘reported infected’ provided in the Greek data. Since the latter number is likely to be smaller than the actual number of infected individuals we have repeated the above considerations using a “virtual time series” formed by magnifying the actual data by a factor, e.g., of 2; it is encouraging for the validity of our conclusions that in this case there was qualitative agreement with the trends discussed above obtained using the available data. Second, our estimates of the effect of easing the lockdown measures are based on the assumption that the characteristics of the virus remain invariant. However, viruses tend to mutate. Thus, both the infectivity and virulence of the virus that caused the ‘first wave’ may change. In this case not only the parameter *c* but other parameters will change.

What are the implications of the above results for policy makers? In this connection, it is important to remark that a definitive connection of the parameter of interest (*ζ)* with non-pharmacological practices such as proper hygiene, wearing gloves and a face-covering mask, social distancing, etc. is presently lacking. As a relevant note of caution, we highlight that *c* does not solely depend on contacts but also on the probability of infection given a contact which is proportional to the viral load (i.e., the viral concentration in the respiratory-tract fluid) of expelled respiratory droplets^36^. Clearly, the above additional protective measures reduce the viral load and therefore *c*. A more systematic experimental determination of the role of such protective measures in the presently modified reality of inter-personal contacts would thus be important towards further assessing safe thresholds of contact increase.

Despite the various limitations of our analysis, the standing connection of *c* within SIR models^41^ with the number of contacts suggests that policy makers should *strongly recommend that people above 40 years of age continue to observe protective measures as strictly as it was done during the full lockdown period*. On the other hand, it appears that the young require less protection. By implementing the above recommendations, it may be possible to allow the society and of course the economy to function, and at the same time to avoid a humanitarian disaster. Consider for example the case of primary schools; members of the teaching stuff younger that 40 years of age could perhaps function normally. On the other hand, members older than 40 years of age would be best served by adhering to lockdown restrictions. Similarly, if the parents of the children are younger than 40, they can perhaps function normally; however, if they are older that 40, they should continue to observe strict protective measures. Moreover, as is self-evident, individuals with medical conditions that place them in high risk, should also follow the stricter measure recommendations applicable to the older population.

Further studies are needed to delineate better the risks associated with different age groups. The results presented here should be considered only as indicative, broad guidelines. Indeed, when reaching the above conclusions, we used the partition of data available in the country of Greece. It is conceivable that the results would not dramatically change if a partition of COVID-19 patients was available for infections and deaths under vs. over the age of 50. Nevertheless, the above analysis should serve as a note of caution that for people over the age of 40 a *substantial* additional degree of caution is needed. We believe that further studies with natural generalizations of the model to a larger number of age groups would be particularly valuable in that regard. Additionally, here we did not attempt to include existing health factors that may affect how prone a sick individual may be towards a fatal result. In a larger scale model such as the one herein, this appears sensible also from the point of view of model identifiability^42^. However, in a smaller number of compartment model examining the potential fate of sick individuals, it would be particularly interesting to assimilate data about their existing medical conditions to more accurately gauge the associated death probability.

Although in this work we have concentrated on Greece, we expect that similar conclusions are also valid for other countries. Actually, taking into consideration that the numbers of deaths and of infected individuals are quite low in Greece in comparison with other countries during the first wave of the COVID-19 pandemic, it follows that the need for considering the above recommendations is even more crucial in these countries. At the same time, the availability of data that would allow the broader applicability of this type of modeling would be of paramount importance in evaluating the course of the pandemic (the corresponding critical value of *c* etc.) in different locations and under different population distributions, climate/weather conditions etc.

In this work the primary composite end point was the number of deaths and the number of infected individuals. A more detailed analysis should examine the number of patients admitted in the intensive care unit, as well as several other consequences of a COVID-19 infection (including, ideally, age-resolved time series for the evolution of hospitalizations and recovered individuals). For example, it is well known that many viral infections can cause a variety of post-viral illnesses, including many neurological conditions, such as the Guillain–Barré syndromes (which comprise a spectrum of polyneuropathies); surprisingly, in the case of COVID-19 such neurological syndromes were not reported until very recently. However,^43,44^ summarizes the documentation of 11 cases of Guillain–Barré syndromes, and warns that other devastating post-viral neurological conditions are also expected to be associated with COVID-19). The possibility of these devastating conditions implies that the recommendations suggested in this work may also bear a broader impact, in connection to such effects.

## Methods

### Discussion of Model Details

For our model we assume that *each* of the two sub-populations has its own individual characteristics, 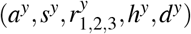 and correspondingly 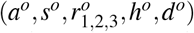. We assume that this is the only way in which the two populations differ. Of course, we identify the dependent variables as pertaining to each of the two populations, i.e., we split the population into (*T*^*y*^, *E*^*y*^, *A*^*y*^, *I*^*y*^, *H*^*y*^, *R*^*y*^, *D*^*y*^) and correspondingly (*T*^*o*^, *E*^*o*^, *A*^*o*^, *I*^*o*^, *H*^*o*^, *R*^*o*^, *D*^*o*^). Then, each of the equations (1)-(5) remains identical as before with the exception that each term in them carries a superscript ^*y*^ or ^*o*^ depending on whether they refer to the young or older population. To avoid cluttering the work with equations, we do not rewrite these equations here.

Instead, we focus on writing the one distinguishing equation, namely the equation for the exposed, because there an additional key assumption needs to be discussed. We now have, instead of (6), two equations for the exposed pertaining to the two populations:

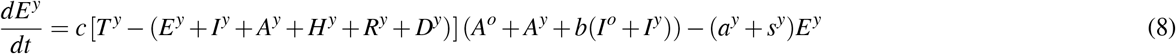

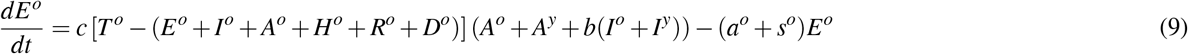

With these equations we have a full dynamical system for the young and older populations. Implicit, however, within this dynamical system is the following modeling assumption: in Eqs. (8)-(9), we have assumed a *single* prefactor *c* and a single coefficient *b*. This assumes that young infectious agents (*A*^*y*^ + *bI*^*y*^) interact with a young susceptible person (first bracket in Eq. (8)) in *exactly the same way* that older infectious agents (*A*^*o*^ + *bI*^*o*^) interact with a young susceptible person. Since it is not clear that this is indeed the case, in a generalized form of the model, one could assume a factor *c*^*yy*^(*A*^*y*^ + *b*^*yy*^*I*^*y*^) for the contact interaction of a young with a young and similarly a different factor *c*^*yo*^(*A*^*o*^ + *b*^*yo*^*I*^*o*^) for the interaction of an older infectious individual (given by this expression) with a young susceptible one. Similarly in this setting, Eq. (9) would include a different mode of interaction between young infectious agents and older susceptibles (proportional to *c*^*oy*^(*A*^*y*^ + *b*^*oy*^*I*^*y*^)), and between older infectious agents and older susceptibles (proportional to *c*^*oo*^(*A*^*o*^ + *b*^*oo*^*I*^*o*^)). We have analyzed this generalized model, which we have called anisotropic. Actually, we obtained slightly better fits for the numbers of infections and deaths using this model. Nevertheless, we opted against showing the results of the latter computations regarding the effect of varying *ζ*. The reason for this decision is that several more parameters are needed for the specification of the anisotropic model; these parameters characterize the ^*yy, yo, oy*^ and ^*oo*^ interactions (i.e., 4 pairs of (*b, c*) parameters instead of 1). This enlarges significantly the space of possible fitting parameters; actually, in this case we have observed the existence of multiple possible local minima (although distinct, these minima yield similarly adequate fitting results). A mathematical formulation of this issue is associated with the notion of identifiability of parameters in such SIR type models^42^. Hence, despite the potential slight improvement of the anisotropic model towards capturing the available data, we feel that the simpler model still provides a quite accurate picture of the impact of the different easing of lockdown strategies.

In the subsection below, we explain how to fit the different parameters of the model of Eqs. (1)-(5) (for both the young and the older sub-populations), as well as (8)-(9), in line with the discussion above regarding the simpler versus the anisotropic model.

### The Optimization Routine

In what follows, we explain the method for obtaining the optimized parameters of the model and also discuss simulation results for the case of Greece. In order to seek the optimized parameters, we look for the minimization, under the constraints imposed by the variation intervals of Tables 1 and 2, by means of the Matlab function fmincon, of the norm

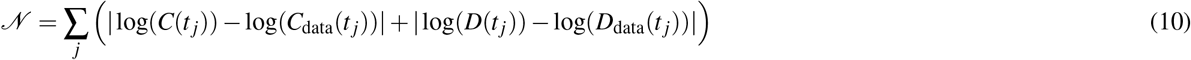

for the single-population version of the model, and

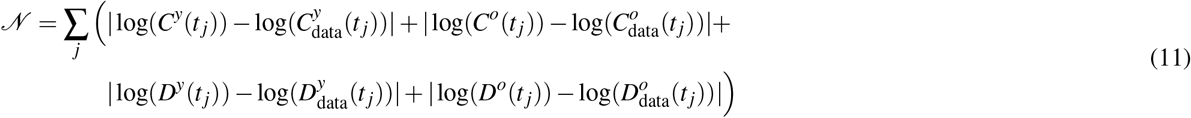

for the the two-population version. *C*(*t*) represents the cumulative infections, which consists of all the people that have passed some symptomatic form of the infection. In this connection we distinguish between the individuals who recovered from the asymptomatic state *R*_*A*_, and those that eventually recovered stemming from the infected population, *R*_*I*_; the total number of recovered individuals is *R* = *R*_*A*_ + *R*_*S*_. The cumulative number of *reported* infected persons is given by *C*(*t*) = *I*(*t*) + *H*(*t*) + *R*_*I*_(*t*) + *D*(*t*). It is this number of cumulative infections computed from the model that we compare to the corresponding number in the official data.

The variable without subscript corresponds to the result obtained from integrating the model equations whereas the subscript *data* refers to the observed data; both variables are tracked at times *t* _*j*_ (that is, every day). The relevant data were obtained from^39^. Moreover, as indicated above, a potential further partition of ages may be beneficial in its own right. Indeed, in the data such a partition does exist between individuals of 0 *—* 17 years, 18 *−* 39 years, 40 *−* 64 years and over 65 years. This renders the relevant data ripe for a consideration towards a 4-age model in the near future. As an additional note worth adding, we have observed that in the above reports the total number of infections does not match the sum of the above 4 age groups. We are not immediately aware for the reason of this discrepancy (possibly the unavailability of age declaration in some of the infection cases). As an example on the May 4th report, the total number of reported infections is indicated as 2632, yet the sum of 104 infections of up to 17 years, 748 between 18 and 39, 1045 between 40 and 64 and 527 above 64 is 2423, leaving 209 infections of unaccounted age. Hopefully, this too will be clarified in future installments of the relevant data. The numbers that we have used pertain to the age partitions in the relevant table of^39^ and thus our numbers add up to less than the total known number of infections.

It is also worth noting as an aside that using the methodology of^35^, it is possible to obtain some groups of parameters for a single-age population model, which are in a similar ballpark as the parameters obtained via minimization and shown in Table 1. These groups of parameters are *R*_2_ = *r*_2_ + *h* = 0.2467, *R*_3_ = *r*_3_ + *d* = 0.1175, *C*_1_ = *ac/*(*hsd*) = 1.5486 *×* 10^*−*5^, *C*_2_ = *bc/*(*hd*) = 6.4947 *×* 10^*−*6^, *r*_1_ = 0.1464 and *F* = *a* + *s* = 0.61.

## Data Availability

All the data are available upon request of whoever ask for it.

## Acknowledgements

ASF acknowledges support from EPSRC in the form of a a Senior Fellowship. PGK and JCM greatly appreciate numerous helpful discussions with Y. Drossinos, Z. Rapti, and G.A. Kevrekidis. This material is based upon work supported by the US National Science Foundation under Grants No. PHY-1602994 and DMS-1809074 (PGK). PGK also acknowledges support from the Leverhulme Trust via a Visiting Fellowship and thanks the Mathematical Institute of the University of Oxford for its hospitality during part of this work.

## Author contributions statement

A.S. Fokas: Conceptualization, Methodology, Formal Analysis, Writing of the Original Draft. J. Cuevas-Maraver: Software, Investigation, Validation, Visualization, Writing of the Original Draft. P.G. Kevrekidis: Methodology, Formal Analysis, Investigation, Writing of the Original Draft.

